# Missed Opportunities for Chronic Kidney Disease Recognition in Irish Primary Care: A Nationwide Electronic Health Record Study

**DOI:** 10.64898/2026.10.27.26366613

**Authors:** Benjamin M. Jacob, Keelan Eyers, Michael Hayes, Harriet Dickinson, Danyao Jin, Aoife Coughlan, Emma Ryan, Benjamin van Hecke, Patrick Redmond, Austin Stack, Donal Sexton, Donal Bailey

## Abstract

**Objectives:** To examine chronic kidney disease (CKD) diagnostic coding in Irish primary care using electronic health records (EHRs).

**Methods:** This retrospective cohort study used EHR data from 592,888 active patients across 66 Irish primary care practices. Adults aged ≥20 years meeting laboratory-based CKD criteria were identified. CKD coding was defined by the presence of an ICD-10 or ICPC-2 diagnosis code. Coding rates were evaluated overall and by CKD stages, comorbidities, and practices.

**Results:** Among 19,382 adults with laboratory-confirmed CKD (median age 76 years; 55.7% female), 14.6% had a documented CKD diagnosis code. Coding increased with disease severity, from 4.8% in stages 1-2 to 19.6% in stages 3-5, and varied by comorbidity burden, with only 10.1% of patients without major cardiometabolic comorbidities coded. Substantial practice-level variation was observed, with over half of practices (34/61) recording coding rates below 10%, while four exceeded 40%. Most patients (85.4%) remained uncoded throughout follow-up.

**Conclusions:** This first nationwide study of CKD coding in Irish primary care network demonstrates substantial under-coding despite laboratory evidence, with marked variation between practices. Improving coding consistency and completeness may enhance CKD recognition, support earlier intervention, and improve patient outcomes.

## Introduction

Chronic kidney disease (CKD) is a progressive condition that is often underdiagnosed, posing a significant health challenge.^1^ In Ireland, prevalence estimates range from 4–5% in the general population to approximately 16% in adults ≥50 years, with most cases occurring in later life.^2,3^ Studies such as TILDA, The Irish Longitudinal Study on Ageing, indicate that up to 98% of individuals with CKD are unaware, reflecting its largely asymptomatic nature in early stages.^3^

Diagnostic coding plays an important role in the recognition and management of chronic diseases in primary care.^4^ Standardised coding systems support consistent documentation, patient identification, follow-up, and healthcare planning.^5^ In the context of CKD, accurate coding may facilitate timely monitoring and evidence-based management, while providing insight into current patterns of disease recognition. Despite the importance of early detection and sustained management of CKD, evidence on coding practices in Irish primary care remains limited, and large-scale assessments have been scarce.^6,7^

This is the first study to examine the diagnostic coding among adults with laboratory-confirmed CKD in a nationwide Irish primary care network. Specifically, we quantified CKD coding rates and explored variation by CKD stages, comorbidities, and primary care practice. By providing a large-scale assessment of CKD coding using community-based electronic health records, this study aims to identify opportunities to improve CKD recognition and inform strategies to support earlier intervention and better patient outcomes.

## Methods

### Study design and data source

This study used routinely collected EHRs from the Clinical Research and Data-Driven Learning Environment (CRADLE) database. CRADLE consolidated data from 66 general practitioner practices within the Centric Health network across Ireland, covering 592,888 active patients in 2025. Adults aged ≥20 years with laboratory-confirmed CKD were identified using EHR data. Coding prevalence was evaluated by quantifying the proportion of patients with a recorded CKD diagnostic coding among those with laboratory-confirmed CKD, defined as two consecutive eGFR values <60 mL/min/1.73 m^2^ at least 90 days apart, or a single uACR value ≥3 mg/mmol. This pragmatic definition was applied in the main analyses, and two alternative definitions of laboratory-confirmed CKD were applied as sensitivity analyses (Supplementary Figure 1, Supplementary Table 1). CKD stages were classified according to the most recent eGFR measurement available at the time of CKD identification. Coded CKD was defined using prespecified ICPC-2 and ICD-10 diagnosis codes recorded in EHRs (Supplementary Table 2).

### Variables and outcomes

Demographic variables included age, sex, laboratory values, and registration status. Comorbidities were identified using harmonised diagnostic codes from the International Classification of Primary Care, Second Edition (ICPC□2), and the International Statistical Classification of Diseases and Related Health Problems, Tenth Revision (ICD□10), for CKD, hypertension, type 2 diabetes mellitus (T2DM), congestive heart failure (CHF), and cardiovascular disease (CVD) (Supplementary Table 2). Laboratory values were standardised across practices, with eGFR expressed in mL/min/1.73 m^2^ and uACR in mg/mmol. The primary outcome was the proportion of patients with laboratory-confirmed CKD who had a recorded CKD diagnosis code. Time to coding was assessed by examining the distribution of the interval between the first laboratory evidence of CKD and the assignment of a formal CKD diagnosis code.

### Bias and quality control

The study population was drawn from Centric Health practices, which may differ from the national population. All patients active on the index date were included to minimise selection bias. Laboratory and diagnostic coding data were extracted directly from electronic records, and missing data were interpreted as reflecting either absence of documentation or incomplete recording in routine clinical practice. Quality control measures included harmonisation of laboratory units and diagnostic codes.

### Statistical analysis

Descriptive statistics were presented as counts and proportions with 95% confidence intervals (CIs) for categorical variables, and as means with standard deviations or medians with interquartile ranges (IQRs) for continuous variables, depending on distribution. Coding rates were summarised overall and by CKD stage, comorbidity subgroup, and primary care practice.

## Results

### Patients Characteristics by Coding Status

Within the active adult patients aged ≥20 years, 19,382 individuals met the criteria for the laboratory-confirmed CKD cohort (Supplementary Figure 1). A total of 2,831 patients with coded CKD were identified.

Among laboratory-confirmed CKD patients, patients with a recorded CKD diagnosis tended to be slightly older than those without a CKD code (median age 78 years [IQR 71-84] vs 75 years [66-82] years) and were more likely to be male (48.6% vs 43.5%) (Supplementary Table 2).

Overall, eGFR testing was common in both coded and uncoded groups (97.7% vs 92.9%), with consistently high testing rates across all CKD stages (>87%), while uACR testing was performed more frequently among uncoded than coded patients (32.9% vs 19.0%). Within both groups, uACR testing rates increased with CKD severity, ranging from 37.8% and 61.8% in stage 1/2 CKD to 30.7% and 21.9% in stage 5 CKD among coded and uncoded patients, respectively (Supplementary Table 3).

### CKD Coding Rate Overall, Across Disease Stages and Primary Care Practices

Overall, the coding rate was 14.6% (14.1–15.1) across all CKD stages. This estimate ranged from 12.5% to 16.6% when alternative definitions of laboratory-confirmed CKD were applied (Supplementary Table 1). Coding rates increased with CKD severity. As shown in Figure 1, 4.8% (95% CI: 4.3–5.3) of patients in stages 1-2 was coded, compared with 17.6% (16.9–18.3) in stage 3, 39.5% (36.4–42.6) in stage 4, and 50.3% (42.3–58.4) in stage 5, and 19.6% in patients with laboratory-confirmed CKD stage 3-5.

**Figure 1.**
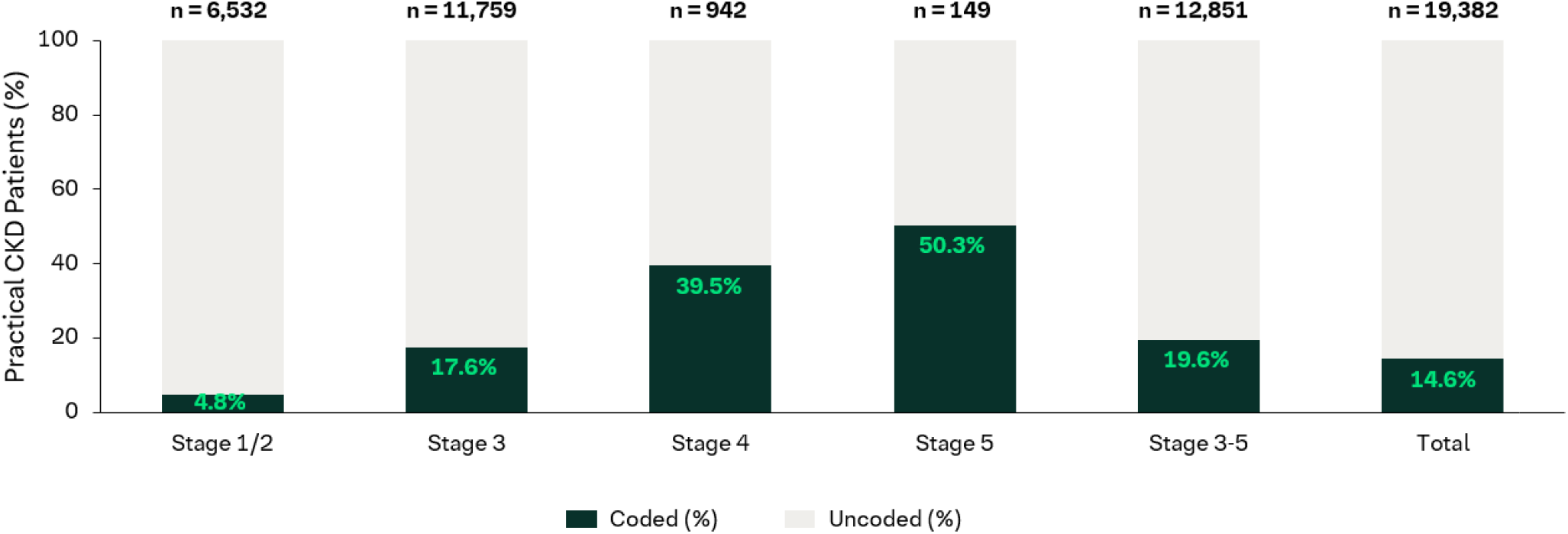
Breakdown of coded and uncoded cases for each CKD stage and total patients with laboratory-confirmed CKD.

### CKD: chronic kidney disease

The prevalence of coded CKD among patients with laboratory-defined CKD varied substantially across practices, ranging from 0.4% to 72.8%, with a median of 8.8% (IQR: 3.9–19.2). Considerable variations were observed across 66 practices: more than half of primary care clinics (34/61) have overall coding rate less than 10%, whereas only four clinics achieved coding rates above 40% (Table 1).

**Table 1.** Coding rate group for laboratory-confirmed CKD patients by clinics.

| Coding Rate | Number of Clinics |
| --- | --- |
| <10% | 34 |
| 10-20% | 12 |
| 20-30% | 4 |
| 30-40% | 7 |
| >40% | 4 |

Note: CKD codes follow ICPC-2 and ICD-10 systems. CKD: chronic kidney disease; ICD: International Classification of Diseases; ICPC: International Classification of Primary Care.

### Time to CKD Coding Overall and by Comorbidities

Among patients with laboratory-defined CKD, the majority (85.5%) were not coded at any point during follow-up. Among coded patients, only a small proportion were coded close to the time of laboratory indication: 37.2% were coded before indication, 24.1% within one year, and 38.6% were coded after more than one year after indication by tests (Table 2).

**Table 2.** CKD coding timing (before, timely, delayed proportion percentages for practical laboratory-confirmed CKD patients), and coding rate by comorbidity.

|  | **Coding rate** | **Coding before CKD indication by tests (%)** | **Coding Timely <= 1 Year after indication by tests (%)** | **Coding Delayed > 1 Year after indication by tests (%)** |
| --- | --- | --- | --- | --- |
| **Overall CKD** | 14.5% | 37.2 | 24.1 | 38.6 |
| **Comorbidity subgroups** |  |  |  |  |
| Hypertension | 19.8% | 45.4 | 23.1 | 31.5 |
| T2DM | 15.2% | 44.7 | 19.4 | 35.9 |
| CHF | 24.6% | 56.6 | 21.3 | 22.1 |
| CVD | 18.5% | 45.0 | 22.7 | 32.4 |
| No Comorbidity from list | 10.1% | 20.4 | 28.1 | 51.5 |

CHF: congestive heart failure; CKD: chronic kidney disease; CVD: cardiovascular disease; T2DM: type 2 diabetes mellitus.
Note: Proportions are expressed as a % of total Practical Laboratory-confirmed CKD patients.

Coding rates varied by comorbidity and were highest among patients with CHF (24.6%), followed by hypertension (19.8%), CVD (18.5%), and T2DM (15.2%), whereas patients without any of the listed comorbidities had the lowest coding rate (10.1%). Across all comorbidity groups, a substantial proportion of CKD diagnostic codes were recorded before laboratory indication, ranging from 20.4% among patients without listed comorbidities to 56.6% among those with CHF. Timely coding within one year of CKD indication by testing occurred in approximately one-fifth to one-quarter of patients (19.4% - 28.1%), while delayed coding beyond one year was common, particularly among patients without listed comorbidities (51.5%), compared with those with hypertension (31.5%), T2DM (35.9%), CVD (32.4%), and CHF (22.1%) (Table 2).

## Discussion

### Main findings

This nationwide study demonstrates substantial under-coding of CKD in Irish primary care. Despite laboratory evidence of disease, fewer than one in six (14.6%) patients with CKD had a recorded diagnosis code, and most (85.5%) remained uncoded throughout follow-up. Although coding rates increased with advancing CKD stage, recognition remained low (19.6%) even among CKD stages 3-5.

Coding varied considerably by both patient characteristics and practice. Patients with cardiometabolic comorbidities were more likely to have CKD coded, whereas those without major comorbidities had the lowest coding rates. Marked practice-level variation was also observed, suggesting inconsistency in CKD recognition and documentation across primary care settings.

Taken together, these findings indicate that CKD remains substantially under-coded in Irish primary care, with diagnosis coding influenced by disease severity, comorbidity burden, and individual practice patterns, representing a missed opportunity for timely identification and management of CKD.

### Comparison with existing literature

Our findings of substantial gaps in CKD coding in Irish primary care were consistent with patterns reported across Europe. In line with our observation that limited uACR testing may underestimate CKD prevalence, a population-based cohort study in France (n = 11,429) found that most CKD cases were identified through increased albuminuria rather than reduced eGFR. This highlighted the importance of routine albuminuria assessment for earlier detection and preventive care, particularly in individuals at higher risk, including those with hypertension, T2DM, and CVD.^8^

CKD prevalence estimates vary widely across Europe, ranging from 3.3% in Norway to 17.3% in northeast Germany for stages 1–5, and from 1.0% in central Italy to 5.9% in northeast Germany for stages 3–5. These differences may reflect variation in detection and monitoring practices. ^9^ Underdiagnosis remains a concern; for example, German data indicated that only 47% of patients meeting laboratory-based CKD criteria had a documented diagnosis.^10^ Together with our findings, this highlights opportunities to improve consistency in CKD coding.

Targeted CKD screening in high-risk adults—those ≥45 years with comorbidities such as T2DM, hypertension, heart failure, or prior myocardial infarction or stroke—could substantially reduce dialysis initiation, cardiovascular events, mortality, and kidney replacement therapy costs, while also lowering the environmental impact of CKD care. These benefits were accompanied by a modest increase in early-stage CKD prevalence, reflecting earlier diagnosis and longer survival. ^5^

These findings indicated that much CKD care may have been delivered through broader cardiometabolic pathways rather than in direct response to CKD, suggesting that earlier coding may have supported more organised care and aligned with the policy rationale of the planned Irish CKD chronic disease management programme; however, where differences were limited, other barriers beyond formal coding alone may have influenced care delivery.

### Limitations

This study had several limitations. The CKD definition relied primarily on eGFR, as uACR testing was infrequent, which limited identification of stage 1-2 CKD and may have resulted in underestimation of coding rates in earlier-stage disease. Coding practices may not have fully reflected clinical recognition due to inconsistencies in ICPC□2 coding and system-related constraints, meaning that some CKD cases may have been recognised but not formally coded. In addition, the broad ICPC-2 code set used to identify CKD was intended to maximise case capture and may have included some kidney-related conditions that are not specific to CKD. Comorbidity analyses relied on coded diagnoses, which may have resulted in under-ascertainment of comorbid conditions and influenced subgroup comparisons. Finally, the findings reflect CKD coding practices within primary care and may not be generalisable to secondary care settings.

### Strengths

To our knowledge, this was the largest CKD study conducted in Ireland and the first to examine CKD coding across a nationwide primary care network. The extensive EHR coverage, with patients contributing several years of data both before and after meeting the study definition of laboratory-confirmed CKD, enhanced the robustness of the analyses and enabled assessment of coding patterns over time. The findings identified opportunities to improve CKD identification among individuals meeting laboratory criteria, thereby supporting earlier detection and more appropriate management.

### Interpretation: implications for research and practice

These findings highlight important opportunities to strengthen CKD recognition in Irish primary care. Improving the consistency and completeness of CKD coding, particularly in earlier disease stages, may support more accurate identification and further management of CKD. The observed variation by comorbidity burden and across practices suggests that CKD recognition is not applied consistently in routine care. Strengthening coding practices, alongside additional support, education, and resources for primary care professionals, may help improve the timely recognition and recording of CKD and support future evaluation of CKD care.

### List of abbreviations

CHF: congestive heart failure; CIs: confidence intervals; CKD: chronic kidney disease; CRADLE: clinical research and data-driven learning environment; CVD: cardiovascular disease; eGFR: estimated glomerular filtration rate, EHRs: electronic health records; GDMT: guideline-directed medical therapy; ICDL10: International Statistical Classification of Diseases and Related Health Problems, Tenth Revision; ICPCL2: International Classification of Primary Care, Second Edition; IQRs: interquartile ranges; TILDA: The Irish Longitudinal Study on Ageing; T2DM: type 2 diabetes mellitus; uACR: urine albumin-to-creatinine ratio

## Data Availability

The datasets analysed during the current study are not publicly available. Access to the data may be available from the corresponding author on reasonable request and with permission from the relevant data custodians, subject to applicable governance and approval processes.

## Declarations

### Ethical approval and consent to participate

The study was approved by the Irish College of General Practitioners (ICGP) Research Ethics Committee (Ref: 2024_2682; 12 December 2024). No additional ethical approval was required.

The requirement for informed consent was waived due to the retrospective nature of the study and the use of anonymised or pseudonymised routinely collected healthcare data. All methods were carried out in accordance with relevant guidelines and regulations.

## Consent for publication

Not applicable as no individual person’s data or images are included in this manuscript.

## Availability of data and materials

The datasets analysed during the current study are not publicly available due to data governance restrictions and patient confidentiality requirements associated with the Clinical Research and Data-Driven Learning Environment (CRADLE) database. Access to the data may be available from the corresponding author on reasonable request and with permission from the relevant data custodians, subject to applicable governance and approval processes.

## Authors’s contribution

The authors meet criteria for authorship as recommended by the International Committee of Medical Journal Editors (ICMJE) and did not receive payment related to the development of the publication. BMJ and KE contributed to the data analysis and interpretation. AS and DS contributed to the clinical interpretation of the results. HD, DJ, AC, ER, BVH, PR, MH, and DB contributed to study conception, design, and oversight. All authors contributed to drafting or critically revising the manuscript and approved the final version.

## Acknowledgments

The authors thank Sarah Cosgrave and Kushagra Rajput (Centric Health IT) and Sam McGlynn and Conor Murphy (ARQ Research Network) for their support of this work. We also extend our gratitude to the staff and patients of Centric Health, whose participation made this research possible. All the authors meet criteria for authorship as recommended by the ICMJE. The authors did not receive payment related to the development of the manuscript. Irene Fernández Meco (BPharm) and David Herrero (PhD) provided medical writing support which was contracted by and funded by Boehringer Ingelheim. Boehringer Ingelheim was given the opportunity to review the manuscript for medical and scientific accuracy as well as intellectual property considerations. The study was funded and supported by Boehringer Ingelheim. The funder contributed to the study design but did not have access to the data and was not involved in the data analyses.

