## Supplemental file for "Missed Opportunities for Chronic Kidney Disease Recognition in Irish Primary Care: A Nationwide Electronic Health Record Study"

**SUPPLEMENTARY MATERIAL**

*Supplementary Figure 1. Inclusion flowchart for the overall study population.*

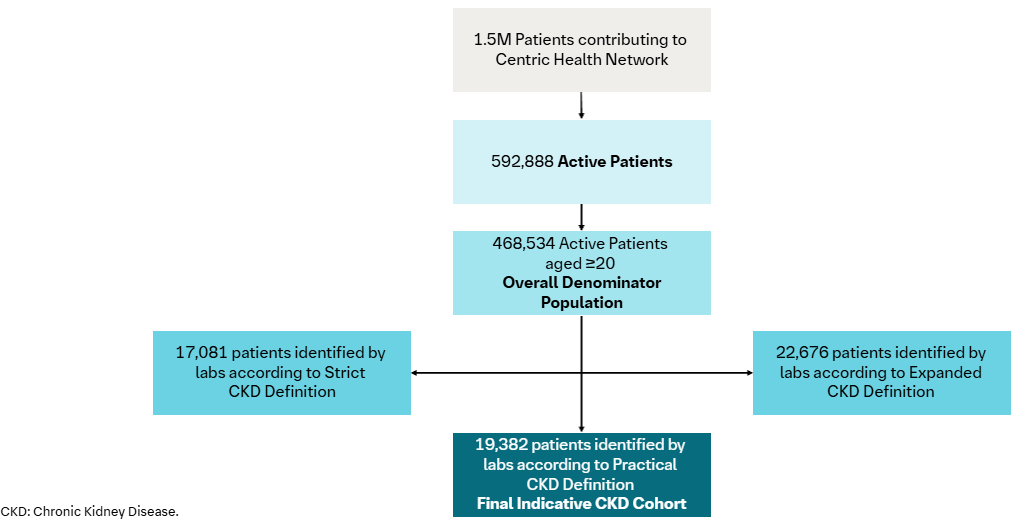

*Supplementary Table 1. Operational definitions of laboratory-confirmed CKD*

| Definition | Purpose | eGFR criterion | uACR criterion |
| --- | --- | --- | --- |
| Strict | Approximate KDIGO-style chronicity-based ascertainment | Two eGFR values <60 recorded ≥90 days apart within 12 months | One uACR >70, or two uACR values 3–70 recorded ≥90 days apart |
| Practical | Primary analysis; reduce under-ascertainment in routine care | As strict definition | As strict definition, or a single uACR >3 |
| Expanded | Sensitivity analysis and comparison with TILDA estimates | As practical definition, or a single most recent eGFR <60 without a prior normal eGFR in the preceding 12 months | As practical definition |

eGFR is expressed in mL/min/1.73m² and uACR in mg/mmol. CKD: chronic kidney disease; eGFR: estimated glomerular filtration rate; uACR: urine albumin-to-creatinine ratio.

*Supplementary Table 2. Code list for coded diseases/comorbidities.*

| Disease | ICD-10 codes | ICPC codes |
| --- | --- | --- |
| CKD | N18, N25, I12, E10.2, E11.2 | U88, U90, U98 |
| Hypertension | I10, I11, I15, I12, I13 | K85, K86, K87 |
| T2DM | E11 | T90 |
| CHF | I50, I42.0, I42.9, I11.0, I25.5, I13.0, I13.2 | K77 |
| CVD | I21, I22, I23, I24, I25, I70, I73, I77, I78, I60, I61, I62, I63, I69.0, I69.1, I69.2, I69.3 | K74, K90, K91, K92, K99 |

CHF: congestive heart failure; CKD: chronic kidney disease; CVD: cardiovascular disease; ICD: International Classification of Diseases; ICPC: International Classification of Primary Care; T2DM: type 2 diabetes mellitus.

*Supplementary Table 3. Summary of characteristics for coded vs uncoded CKD in laboratory-confirmed CKD patients.*

| **Characteristic** | **Coded** | **Uncoded** |
| --- | --- | --- |
| Total Number | 2831 | 16551 |
| Age (mean) | 76.3 | 72.8 |
| Age (median; IQR) | 78 (71 – 84) | 75 (66-82) |
| Male (%) | 1375 (48.6%) | 7205 (43.5%) |
| eGFR tested (%)* | 2766 (97.7%) | 15386 (92.9%) |
| Stage 1/2 (%)^†^ | 305 (97.8%) | 5457 (87.6%) |
| Stage 3 (%)^†^ | 2034 (98.2%) | 9332 (96.4%) |
| Stage 4 (%)^†^ | 357 (95.9%) | 527 (92.9%) |
| Stage 5 (%)^†^ | 70 (93.3%) | 70 (95.9%) |
| uACR tested (%)* | 539 (19.0%) | 5460 (32.9%) |
| Stage 1/2 (%)^†^ | 118 (37.8%) | 3850 (61.8%) |
| Stage 3 (%)^†^ | 317 (15.3%) | 1484 (15.3%) |
| Stage 4 (%)^†^ | 81 (21.8%) | 111 (19.6%) |
| Stage 5 (%)^†^ | 23 (30.7%) | 16 (21.9%) |

*Proportion is expressed as a % relative to the total number in the coded/uncoded group. ^†^Proportion is expressed as a % relative to the total number in CKD stage for the coded/uncoded group. CKD: chronic kidney disease; IQR: interquartile range.
